# The effects of community treatment orders (CTOs) on readmission to hospital using electronic health records

**DOI:** 10.1101/2023.11.27.23298057

**Authors:** Justin C Yang, Johan H Thygesen, Nomi Werbeloff, David Kelsey, Dominique Merlande, Joseph Hayes, David PJ Osborn

## Abstract

**Background:** Community treatment orders (CTOs) are used to manage community-based care for individuals with severe mental health conditions who have been discharged from inpatient care. Evidence examining whether CTOs are successful at reducing rehospitalisation has been mixed.

**Methods:** Using deidentified electronic health records from 2009-21, we compared patients who had ever been placed on a CTO (n=836) and two other groups of patients who had never been placed on CTO: patients admitted under Section 3 of the Mental Health Act (n=1,182) and outpatients with severe mental health issues (n=7,651). We examined the association between CTOs and rehospitalisation using within-individual stratified multivariable Cox regression.

**Results:** Patients on CTO were more likely to be male, single, of Black or Mixed ethnicity, and have a severe mental illness diagnosis than patients in the comparison groups. Time spent on CTO was associated with a lower risk of hospitalisation compared to time spent off CTO for the same individual (HR 0.60; 95% CI 0.56-0.64). This decreased risk of hospitalisation remained when we restricted analysis to individuals with a single CTO episode (HR 0.05; 95% CI 0.02-0.11) and when we restricted follow-up time to a patient’s first CTO episode (HR 0.20; 95% CI 0.17-0.25). However, there was no difference in re-hospitalisations when we observed patients starting from the first CTO (HR 1.07; 95% CI 1.00-1.16).

**Conclusions:** We found that patients on CTO were at lower risk of hospitalisation, though this pattern was not observed when we excluded time prior to the first CTO. Further research should consider whether CTOs provide genuine clinical benefit.

## Introduction

Community Treatment Orders (CTOs) are a legal mechanism used in England and Wales to manage individuals with severe mental health conditions who have been discharged from inpatient care ^1^. They are intended to provide support and treatment to individuals in the community, with the goal of preventing relapse and hospital readmission. CTOs can be used for individuals with conditions such as schizophrenia, bipolar disorder, and severe depression who have a history of non-adherence to treatment and/or frequent hospitalisation^1^.

Under a CTO, an individual is required to comply with a specified treatment plan, which may include medication, therapy, and other support services ^2^. They may also be required to attend regular reviews with a mental health professional and/or comply with other conditions such as living at a specific address, abstaining from illicit substances, or not leaving a certain geographical area ^2^. They are also subject to being recalled to hospital under a CTO ^2^. CTOs are typically issued on the recommendation of the Responsible Clinician and an Approved Mental Health Professional and can be in place for up to six months, with the possibility of renewal ^2^. The individual has a right to appeal the decision and the patient may be discharged from a CTO by a the tribunal if the individual’s condition improves or they no longer meet the legal criteria for the CTO ^2^. CTOs are a type of compulsory community treatment (CCT), often used to treat patients with serious mental illnesses, and have been used in Australia, Canada, Israel, New Zealand, and the United States ^3^.

Critics of CTOs argue that they can be used as a form of involuntary treatment and may restrict the individual’s rights and freedom ^4^. Supporters argue that they can be an effective way to provide continuity of care and support for individuals with severe mental health conditions who may be at risk of hospitalization or harm to themselves or others ^5^. Moreover, there is evidence to suggest that there may be disparities in the application of CTOs on certain patient populations ^6^. Some research has highlighted higher rates of CTO use among patients from Black, Asian, and minority ethnic (BAME) backgrounds ^7,8^. These disparities may be due to a combination of factors, including bias and discrimination within the mental health system, cultural differences in understandings of mental health and treatment, and a lack of culturally appropriate services and support ^9^ris.

In part due to these concerns, there has been an interest in understanding whether or not CCTs such as CTOs actually reduce hospital admissions. A systematic review and meta-analysis of randomised controlled clinical trials (RCTs) and observational studies concluded that there was little evidence for CCT in reducing hospital admission length or frequency ^10^, finding no significant reduction in readmission to hospital ^10^ in experimental studies. Some observational studies showed improvements in outcomes among patients on CCT but did not compare these findings with a control group ^10^. Relatively few studies have examined CTOs in the UK. The only RCT in the UK to assess the effectiveness of CTOs, the OCTET trial, found no effect on admission length and frequency for any subgroup at 12 and 36 months ^11,12^. However, criticism exists of the representativeness of patients within this RCT^13^.

The use of electronic health records (EHRs) provides an opportunity to understand disparities in the application of CTOs by providing a comprehensive and detailed view of an individual’s health history, follow-up time, treatment, and outcomes. EHRs can be used to analyse data on the demographic characteristics of individuals subject to CTOs, including race, ethnicity, gender, and socioeconomic status. We aimed to use de-identified clinical data to evaluate patterns and trends in the use and outcomes of CTOs in reducing hospital admissions across different patient populations over extended periods of time. We also aimed to characterise patients who are placed on CTO compared with patients with a similarly severe symptomatology who are not placed on a CTO.

## Methods

### Setting and participants

We used the Camden & Islington NHS Foundation Trust (C&I) Clinical Record Interactive Search (CRIS) database of deidentified electronic health records for this study. C&I is a secondary mental health trust providing mental health, learning disability, and community health services to approximately 470,000 residents in the London boroughs of Camden and Islington. The geographical area is known for its diverse communities, with a high proportion of immigrants and people from ethnic minority backgrounds. Both boroughs have a higher proportion of people in the 20-29 age group as well as relatively high levels of deprivation compared to the London average.

The C&I CRIS database allows for analysis of anonymised EHR data of over 160,000 patients under the care of C&I from 2008 onwards ^14,15^.

### Exposures

We defined our study cohort as any individual aged 16 and above receiving care at C&I who was ever on a CTO at any point in the 13-year period between 1 January 2009 to 31 December 2021. Individuals were followed up from 1 January 2009 or the first recorded date of contact with the Trust, whichever was later, until 31 December 2021 or date of death, whichever was earlier.

We compared this cohort with two comparison groups who were never on CTO: (1) individuals who had been admitted for inpatient care under Section 3 of the *Mental Health Act (1983)* during the study period and; (2) individuals with serious mental illness (defined as having at least one Health of the Nation Outcome Scales (HONOS) ^16^ score in the top 25% of those recorded in C&I CRIS) who were never admitted as inpatients during the study period.

### Outcomes

We calculated the number of episodes and days each patient spent on CTO during the study follow-up period among patients who had at least one recorded CTO episode and the number of episodes and days spent in inpatient care during the study period among patients who had at least one recorded CTO episode or at least one recorded episode under Section 3 of the Mental Health Act.

### Covariates

Sociodemographic variables, specifically, gender, marital status, ethnicity, and area-level index of multiple deprivation, were extracted from CRIS. The index of multiple deprivation (IMD) is a composite measure of relative deprivation for a specific area. We used IMD ranks for lower super output areas (LSOAs), a small geographic area with a population of about 1,500, to calculate tertiles representing high, medium, and low relative area-level deprivation, indexed across LSOAs in England.

In addition, we identified whether or not patients had a history of severe mental illness by identifying patients who had a recorded ICD-10 diagnosis of F20-29 (schizophrenia), F30-1 (manic episode and bipolar disorder), or F32.3/ F33.3 (major depressive disorder with psychotic symptoms).

### Ethics Approval

Ethical approval for use of the C&I CRIS database was obtained from the East of England - Cambridge Central Research Ethics Committee (19/EE/0210) and the project was approved by the C&I Research Oversight Committee.

### Statistical analysis

Firstly, we compared the sociodemographic and clinical characteristics of the main CTO study cohort with the two comparison groups not on CTOs.

For the main analysis of patients who had been placed on a CTO, we performed a stratified Cox proportional hazards analysis comparing hospitalisations during periods on and off CTO. All analyses were stratified by individual patients and adjusted for time-varying confounders, namely: calendar year, age and count of inpatient admissions within the study period. Time spent as an inpatient was omitted to avoid immortal time bias, because patients could not experience a new admission whilst an inpatient. Figure 1 depicts an example of how our stratified Cox proportional hazards regression examined periods on and off CTO for an individual patient, where yellow indicates a period during which a patient was on a CTO and red indicates a period spent as an inpatient (which was excluded from our analysis).

**Figure 1:**
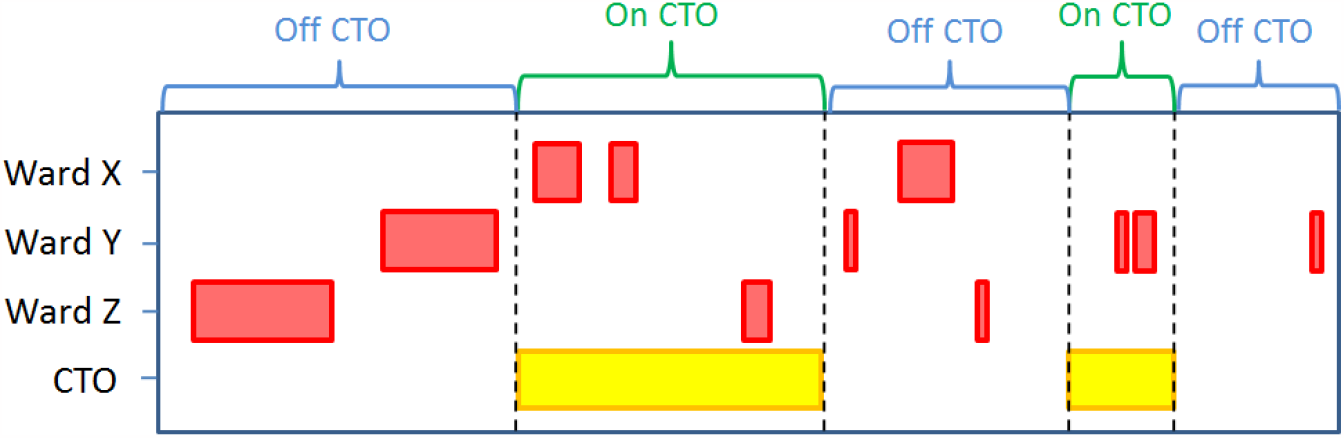
Diagram illustrating periods of hospitalisation coinciding with periods on or off community treatment orders (CTO), of a hypothetical patient.

We conducted three further sensitivity analyses to test whether or not findings from our main analysis held even when specific parameters or assumptions were changed:

1. We restricted the sample to only individuals with <u>one recorded CTO</u> to account for potential bias if there were a systematic difference between individuals who are only ever placed on one CTO from individuals who are repeatedly placed on CTO during their follow up period;
2. We restricted the observation period <u>up to and including the first CTO</u> to investigate the potential that being placed on one CTO and the outcome from that CTO may affect a clinician’s decision to place someone on subsequent CTO episodes, thereby introducing a systematic difference between people who are only ever placed on one CTO and those who are placed on multiple CTOs; and
3. We began observation <u>starting at the first CTO</u> to avoid potential bias introduced by medical care prior to the first CTO in case potential confounders, especially time-varying confounders, played a role in the time leading up to an individual being placed on their first CTO.

Analyses were conducted using R 4.2.4.

## Results

### Characteristics of Patients Placed on CTO Compared with Patients Who Were Not

Sample characteristics are shown in Table 1. We included deidentified data from 836 individuals who were treated under CTO during the study period with a median of 1 (IQR 1-3) CTO episode and a median of 490 (183.0 to 994.0) days spent on CTO. In the comparison groups who were not exposed to CTO, we included 1,182 individuals who were treated under Section 3 of the MHA, and 7,651 severely unwell individuals who exclusively received care in the community. Individuals receiving a CTO were more likely to be male, single, from Black or Mixed ethnic groups, and have a diagnosis of severe mental illness compared to individuals treated on Section 3 or severely unwell outpatients. Compared with individuals who had been treated on Section 3, individuals on CTO had more previous inpatient episodes and had previously spent more days under inpatient care.

**Table 1:**
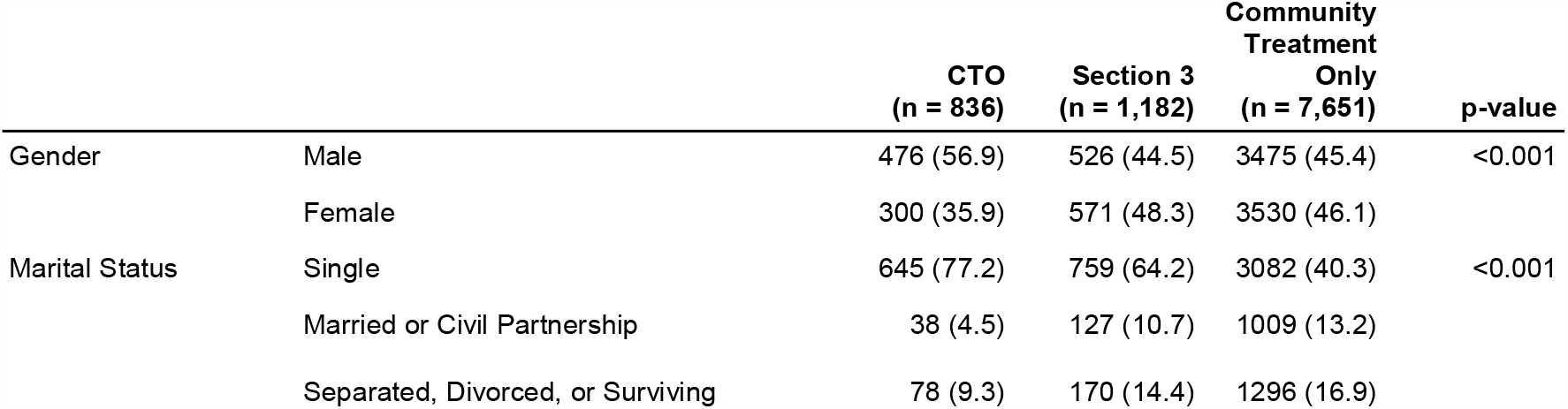
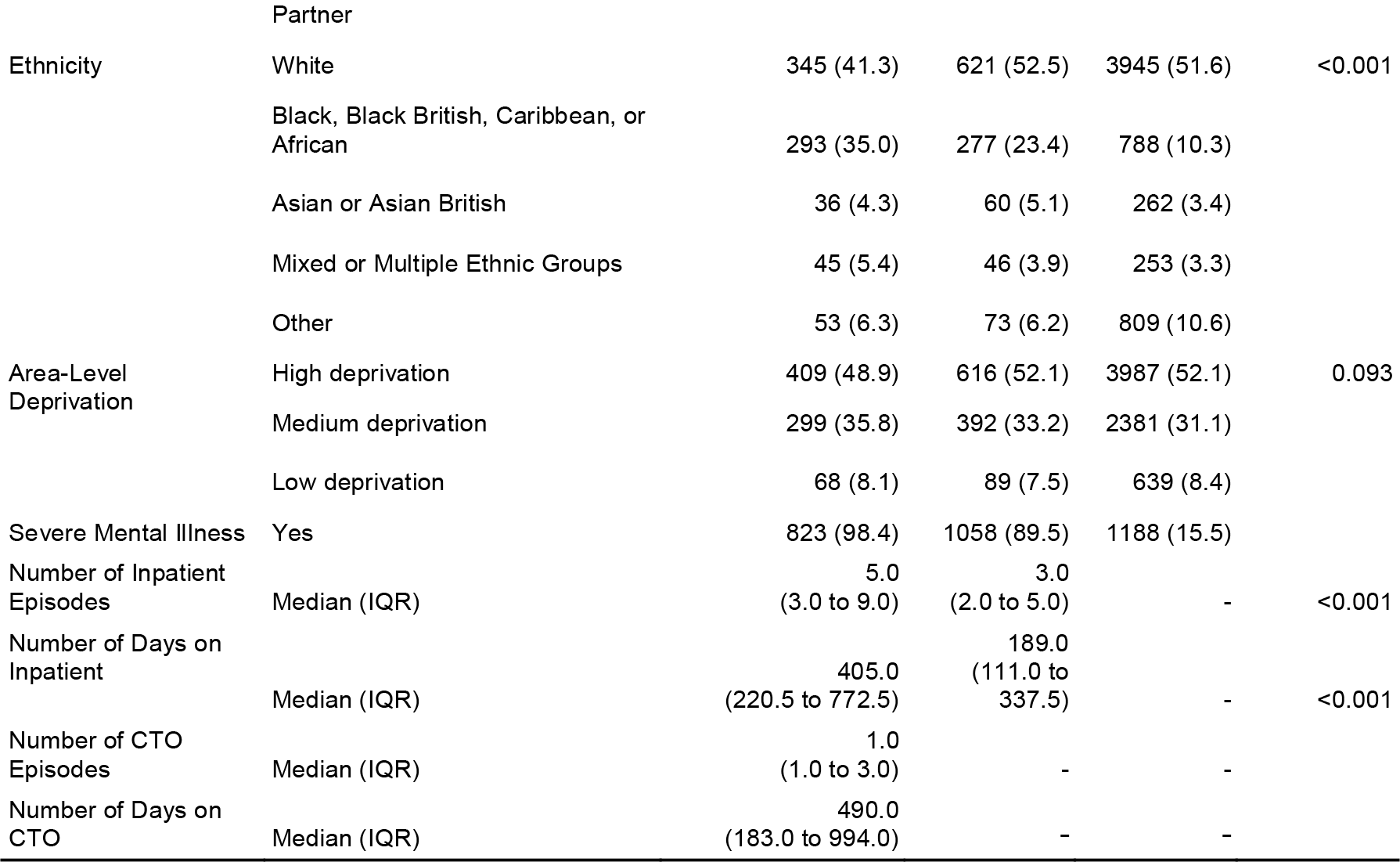
Sample demographics, stratified by intervention. Numbers in (parentheses) are percentages. p-values are provided for Fisher’s exact test.

### Comparison of hospitalisation rates on and off CTO

In our main analysis of individuals with at least one CTO during the study period, we found that time spent on CTO was associated with a lower risk of hospitalisation than time spent off CTO (HR 0.60; 95% CI 0.56-0.64) (Table 2). This decreased risk of hospitalisation was also observed when we restricted analysis to individuals with only one CTO episode (HR 0.05; 95% CI 0.02-0.11) and when we restricted analysis to time up to and including the first CTO (HR 0.20; 95% CI 0.17-0.25). However, we did not observe a difference in risk of hospitalisation when we restricted our analysis to observing patients starting from the first CTO (HR 1.07; 95% CI 1.00-1.16). Results were largely similar between unadjusted univariable regression and adjusted multivariable regression except when we examined individuals starting from their first CTO, where we observed a higher risk of hospitalisation during CTO in both the unadjusted and adjusted models (though, in either case, these were not statistically significant differences).

**Table 2:** Cox proportional hazard results of analysis of risk for hospitalisation.

| Sample | Number of CTOs | Observation Start | Observation End | n | Person-Days Observed | Hazard Ratio (Univariable) | Hazard Ratio (Multivariable*) |
| --- | --- | --- | --- | --- | --- | --- | --- |
| Individuals with at least one CTO | ≥1 | 1 Jan 2009 or first contact with C&I | 31 Dec 2021 or death | 834 | 12,662 | 0.61 (0.57-0.64, p<0.001) | 0.60 (0.56-0.64, p<0.001) |
| Individuals with only one CTO | 1 | 1 Jan 2009 or first contact with C&I | 31 Dec 2021 or death | 262 | 2,612 | 0.04 (0.02-0.09, p<0.001) | 0.05 (0.02-0.11, p<0.001) |
| Individuals up to and including first CTO | ≥1 | 1 Jan 2009 or first contact with C&I | End of first CTO | 834 | 7,256 | 0.30 (0.26-0.36, p<0.001) | 0.20 (0.17-0.25, p<0.001) |
| Individuals starting from first CTO | ≥1 | Start of first CTO | 31 Dec 2021 or death | 834 | 7,336 | 1.02 (0.95-1.09, p=0.584) | 1.07 (1.00-1.16, p=0.062) |

\*Adjusted for year, number of inpatient episodes, gender, marital status, ethnicity, IMD tertile, and diagnosis of severe mental illness

## Discussion

To our knowledge, this is the first study to examine effectiveness of CTOs in reducing rehospitalisation with long follow-up periods, compared to existing literature, in a representative sample. Using anonymised EHR data, we were able to highlight differences in the types of patients most likely to be placed on CTO as well as effects of CTOs on readmission over thirteen years in the second largest sample examined in the UK to date. By comparison, Taylor et al. examined 1,558 individuals followed up over 12 months ^17^ and Vergunst et al. looked at 114 patients over 48 months ^18^.

We observed that patients treated under CTO were more likely to be male, single, of Black or mixed ethnicity, and have a diagnosis of severe mental illness compared with patients treated on Section 3 or severely unwell outpatients in secondary care. The patients who received CTOs were also more likely to have previous inpatient episodes and to have spent more time under inpatient care in the past. In the main analyses, we found that periods on CTO were associated with a lower risk of re-hospitalisation compared to periods off CTO among individuals with at least one CTO. This is consistent with prior findings. This finding remained when we examined only individuals with a single CTO episode or when limiting the follow-up period so it only included time before and during the first CTO episode. However, when we restricted observations to commence from the date of the first CTO, there was no difference in rates of admission whether on or off CTO. Notwithstanding, there is some evidence to suggest that CTOs may reduce the risk of rehospitalisation, especially among those who only ever have a single CTO episode.

### Comparison with other studies

An estimated 5,000 individuals are on CTOs in the UK annually, much more than envisaged when they were introduced and; controversially, those of Black or Black British ethnicity are up to eight times more likely than White people to be placed on CTO ^6^. Our findings showed this national pattern at the local level. As well, our findings were consistent with other studies showing that people placed on CTO were more likely to be unmarried, have a primary diagnosis of psychotic illness, and have a history of longer inpatient admissions ^6^, possibly reflective of increased disease severity among people who are considered for and put on CTOs compared with patients who are treated by other means.

Comparisons with studies set outside the UK are challenging, given major differences in legislation which provide the legal mechanism for CCTs, health care provision, and demography. There are few previous UK-based observational studies which tend to be limited by relatively short follow-up periods and small sample sizes, or are subject to unmeasured confounding ^10^. These studies find that CTOs offer clinical benefits, yet the only UK-based RCT on CTOs failed to find any improvement from CTOs ^11,12^.

### Strengths and limitations

Our study addresses many of the limitations which apply to RCTs of CTOs in terms of generalisability due to strict inclusion criteria. Methodological limitations have been described in non-randomised studies ^10^, such as unmeasured confounding or channelling bias, however we addressed some of these through our within-individual design, by including multiple hospital admissions and performing a number of sensitivity analyses; particularly in excluding the period just prior to the initiation of the first CTO. Furthermore, relatively few studies have assessed follow-up beyond one or two years, while we were able to follow up for over 10 years in some cases, and most published studies have, to the best of our knowledge, only included single CCT episodes.

Yet, limitations remain in our design. There may be important time-varying confounders, such as the availability of social or community support as well as periods of illness stability which cannot be captured from the EHR. In addition, this study only includes data from one mental health service where decisions about CTOs may be relatively homogenous or influenced by a small number of clinicians and may not reflect wider patterns in the use of CTOs across the UK as a whole.

## Conclusions

CTOs, introduced in England over fifteen years ago, continue to be used to treat patients with severe mental illness. Despite a review of the Mental Health Act which recommended a reduction in their use, the use of CTOs has not decreased ^20^, despite concerns about ethnic disparities in the use of CTOs ^19^. They provide a legal mechanism by which patients may be discharged into the community but still be compelled to return to hospital if a clinician deems it necessary. Nevertheless, their potentially coercive nature necessitates greater scrutiny of whether they truly benefit patients or not, and whether there are significant disparities in the use of CTOs based on demographic characteristics.

Our findings highlight the need to further examine whether CTOs truly provide clinical benefits beyond proxy measures of re-hospitalisation and lengths of stay, given patients’ concerns about coercion and potential disparities in their use, and for whom these benefits might apply.

## Data Availability

The data that support the findings of this study are available to approved researchers through Camden & Islington NHS Foundation Trust. The data are not publicly available due to restrictions on data access by the Data Controller in the interest of patient confidentiality.

https://www.candi.nhs.uk/health-professionals/research/ci-research-database

